# Virtual Home Visits Reduce Asthma Burden in Low-Income, Black Communities amidst the COVID-19 Pandemic

**DOI:** 10.1101/2022.11.05.22281903

**Authors:** Arundhati Bakshi, Elora Apantaku, Tracy Marquette, Colette Jacob, S. Amanda Dumas, Kate Friedman, Kathleen Aubin, Shannon Soileau, Shaun Kemmerly

## Abstract

**Objectives:** Pilot an asthma virtual home visits program, and evaluate its reach and ability to promote asthma self-management strategies in underserved communities.

**Methods:** Participants were continuously recruited into the 3-month-long program starting March 2021 and provided with materials related to asthma education. Participants reporting poorly-controlled asthma and home-based triggers were also offered three virtual home visits with a respiratory therapist. All participants were asked to complete a pre- and post-intervention knowledge test and Asthma Control Test (ACT), and a final survey assessing perceptions regarding asthma management and environmental trigger reduction.

**Results:** As of October 2022, 147 participants were enrolled, and 52 had consented and received at least one virtual home visit. Approximately 77% of virtual visit recipients were children, 76% were non-Hispanic Black persons, and 90% were from ‘extremely low’ or ‘low’ income families. Asthma symptoms improved across the whole group, with a median increase of 2.39 points on the ACT score. Knowledge tests revealed that 86% of participants learned about at least one new asthma trigger, with a larger fraction of virtual visit recipients (68% vs. 36% non-recipients) showing an improved score post-intervention. About 75% of participants reported feeling more empowered to self-manage their asthma after participating in the program, and reported a significant improvement in their quality of life due to asthma.

**Conclusions:** The program successfully provided virtual asthma education to underserved, at-risk communities, and improved asthma outcomes for participants. Similar virtual models can be used to promote health equity, especially in areas with limited access to healthcare.

**Summary Box:** *What is the current understanding on this subject?:* Home-based interventions are known to be beneficial for improving asthma outcomes, especially among children; however, in-person visits have been a challenge during the COVID-19 pandemic.

*What does this report add to the literature?:* This report suggests that virtual, home-based models for asthma education are a viable alternative to in-person visits. As such, they can be an important tool for promoting health equity, especially in areas with limited access to healthcare.

*What are the implications for public health practice?:* Public health practitioners should be educated regarding the benefits of home-based asthma interventions, including virtual programs, as an adjuvant to standard clinical practices.

## Introduction

Asthma is a common chronic disease in the United States (US), that disproportionately affects Black and low-income communities, and can lead to hospitalization among children and adolescents.^1^ It can cause lifelong disability and has both genetic and environmental risk factors.^2^ Additionally, environmental triggers present in places where people with asthma spend most of their time can trigger exacerbations limiting meaningful activities and impairing quality of life (QOL). The US Environmental Protection Agency (EPA) estimates that Americans spend 90% of their time indoors; thus the place where environmental triggers are most likely to worsen asthma symptoms is typically the patient’s home.^3^ Traditional clinic-based management of asthma that focuses only on appropriate medication use limits the amount of education patients receive about potential triggers in their own home.

Underserved communities face additional risk factors as lower quality homes have higher exposures of common household triggers.^4^ Asthma disproportionately affects children from Black and low-income families^1,2^, and low-income families have been shown to live in homes with worse air quality, poor moisture control, and more allergens and dust.^4^ Additionally, renters are more likely to be exposed to smoke and mold in their homes and are more likely to visit the Emergency Room (ER) or urgent care facilities for asthma symptoms than those who can afford to own their own homes.^5^ Thus, discussion of clinical management at the doctor’s office, while important, may miss important home and lifestyle modifications that can improve symptoms, especially within at-risk populations.

Home-based interventions that promote both clinical and environmental management of asthma have been shown to be beneficial.^6,7^ However, in-person home visits can have geographic limitations based on availability of care providers, especially in rural areas of states like Louisiana where healthcare access is limited. Moreover, during the COVID-19 pandemic, in-person home visits also carried risk of disease transmission for both patients as well as health care workers.

Having identified a need to improve asthma outcomes in underserved communities on the cusp of the pandemic^8,9^, the Louisiana Department of Health (LDH) attempted to address these issues collectively by piloting a virtual model for asthma home visits as part of their BREATHE program (<u>B</u>ringing <u>R</u>espiratory Health <u>E</u>quity for <u>A</u>sthmatics <u>T</u>hrough <u>H</u>ealthier <u>E</u>nvironments). This Virtual Home Visits (VHV) program was intended to serve Louisiana residents with poorly-controlled asthma and significant home environmental concerns, with a three-fold objective of improving patients’ asthma control, disseminating knowledge of environmental asthma triggers and empowering people to self-manage their asthma, such that the impact of asthma on their QOL would be reduced.

### Program Description

The BREATHE VHV pilot program **(Figure 1)** was designed to bring asthma and Healthy Homes education to assist with asthma management in areas that have a high burden of asthma, COVID-19, and social and environmental vulnerability.^10^ Louisiana residents interested in participating were identified through COVID-19 contact tracing, clinic referrals and direct community outreach. Interested individuals were screened for enrollment with questions that determined the level of asthma control and exposure to home environmental triggers **(Table 1A-B)**, and the data stored in a custom REDCap data management system. Individuals with a score of ≥2 for asthma severity as well as home environmental concerns were eligible for up to three VHV, one month apart. VHV were conducted on the telehealth platform, ANDOR, by a respiratory therapist (RT) at Our Lady of the Lake Children’s Hospital (OLOLCH). All enrollees, regardless of VHV eligibility, were provided educational materials via postal or electronic mail, which included information about both the clinical and environmental management of asthma. All educational materials can be accessed on the LDH BREATHE website (https://ldh.la.gov/page/BREATHE). The first VHV was focused on building rapport with the patient, and discussing asthma management tools, medication adherence, and simple home remedial tips, such as using mattress covers, vacuuming, and asthma friendly cleaning agents **(Supplement 1)**. The remaining two visits were designed to receive updates on patients’ asthma status, discuss takeaways and reinforce lessons from previous visits. After approximately three months, participants were administered a final survey **(Supplement 2)** measuring their attitudes and perceptions regarding asthma management and environmental trigger reduction. Participants were also asked to complete a pre- and post-intervention knowledge test **(Supplement 3)** and age-appropriate Asthma Control Tests (ACT; **Supplement 4**).^11^ For child participants under age 18 years, a parent or guardian completed all surveys, except questions on the ACT that required the child to answer them. These data were then analyzed to test the efficacy of the pilot program.

**Table 1.** Asthma-related clinical and environmental characteristics of all BREATHE patients and demographic characteristics of VHV recipients.

| **A) Clinical Characteristics of ALL participants (n=147):** | **N (% All)** |
| --- | --- |
| used rescue inhaler $\geq 2$ times in a typical week (*score=1*) | 101 (69%) |
| woke at night $\geq 2$ times in a typical week with asthma symptoms or cough (*score=1*) | 87 (59%) |
| filled their rescue medicine $\geq 2$ times in a year (*score=1*) | 90 (61%) |
| visited the ER $\geq 2$ times and/or were hospitalized $\geq 1$ time in the last 6 months (*score=2*) | 59 (40%) |
| suffer from allergies (*score=1*) | 118 (80%) |
| **B) Home Environmental Characteristics of ALL participants (n=147):** | **N (% All)** |
| had moderate or heavy dust build-up in their home (*score=1 for moderate; 2 for heavy*) | 69 (47%) |
| had seen mold, smelled musty odors or experienced water leaks (*score=1*) | 40 (27%) |
| had problems with pests in the last 3 months (*score=2*) | 44 (30%) |
| had someone who smoked in the home in the past 7 days (*score=2*) | 13 (9%) |
| had gas cooktop (*score=1*) | 46 (31%) |
| did not use an exhaust fan or open a window when cooking on the stove (*score=1*) | 40 (27%) |
| have furry or feathered pets in the home (*score=1*) | 60 (41%) |
| used cleaning agents or perfumed products that have a strong odor (*score=1*) | 116 (79%) |
| **C) Demographic Characteristics of VHV recipients (n=52):** | **N (% VHV)** |
| Age group: Children (0-17 years of age) | 40 (77%) |
| Age group: Adults (18-64 years of age) | 11 (21%) |
| Age group: Older adults (65+ years of age) | 11 (2%) |
| Sex: Female | 29 (56%) |
| Race: Black | 40 (76%) |
| Race: White | 12 (24%) |
| Homeownership: Renters | 32 (62%) |
| Income level: Extreme low (<30% AMI) | 21 (41%) |
| Income level: Low (<50% of AMI) | 25 (49%) |
| Income Level: Moderate (<80% AMI) | 5 (10%) |
| Top county (parish) of residence: East Baton Rouge | 23 (44%) |

ER = Emergency Room
VHV = Virtual Home Visit
AMI = Annual Median Income (US Housing and Urban Development). Designations based on AMI for a family of four (average family size of participants) in East Baton Rouge Metropolitan area (where majority of participants resided).

**Figure 1:**
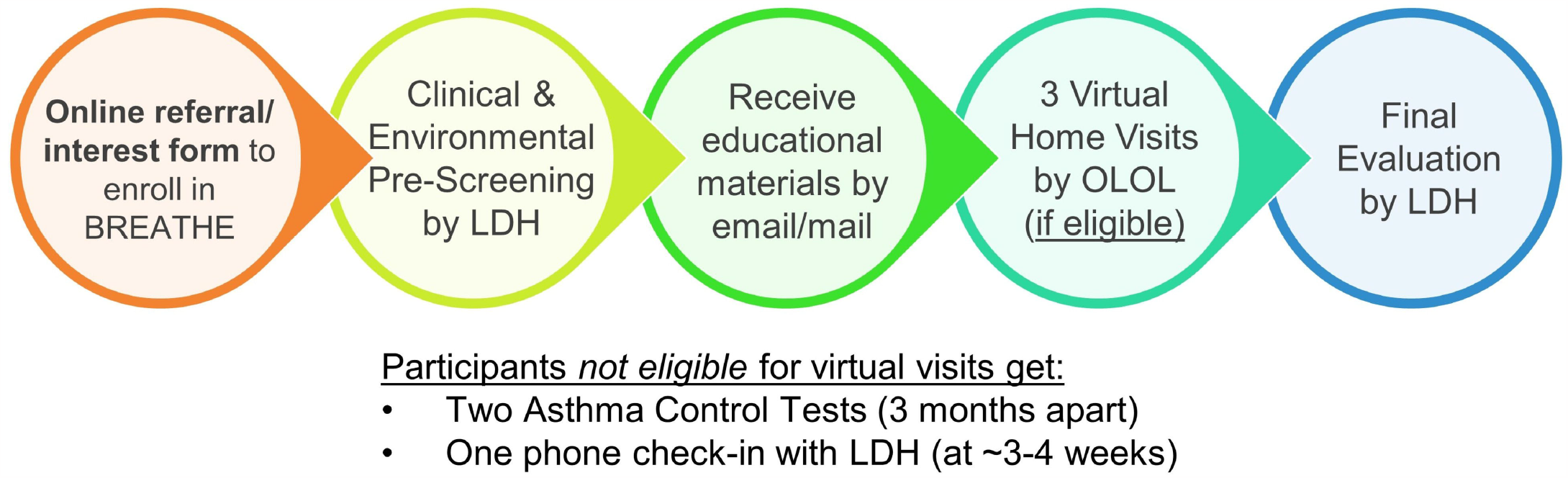
REATHE Program Design. All interested patients were enrolled via phone or an online interest form (www.ldh.la.gov/breathe-enroll). Based on the clinical and environmental pre-screening information they provided, patients were offered three virtual home visits by a RT. After three months, participants were evaluated on their asthma status as well as attitudes and perceptions towards asthma management after participation in the program.

### Evaluation Purpose and Criteria

The program evaluation was designed to assess four focus areas:

1. <u>Populations and areas served</u>; measured by demographic characteristics of VHV recipients. The VHV aimed to serve children, Black and low-income patients residing in previously identified areas of concern^9^, namely East Baton Rouge (EBR), Caddo, Jefferson, and Orleans Parishes (counties), the latter two together comprising the Greater New Orleans (GNO) Area.
2. <u>Asthma symptom control</u>; measured by pre-vs. post-intervention ACT scores. Success was defined as final ACT scores being higher than initial ones, with at least 50% patients showing improvement. We hypothesized that patients who received the VHV would show a greater improvement than those who did not.
3. <u>Knowledge about environmental trigger management</u>; measured by pre-vs. post-intervention knowledge test scores. Success was defined as at least 50% patients learning about a minimum of one environmental asthma trigger and improving their post-test score. We hypothesized that patients who received the VHV would show a greater improvement in their knowledge of environmental trigger reduction than those who did not.
4. <u>Attitudes and perceptions about asthma self-management and impact of asthma on quality of life</u>; gauged using a post-intervention survey asking about their perceptions regarding asthma symptoms, asthma education, environmental trigger reduction and sense of empowerment regarding asthma self-management after participation in the program. Success was defined as at least 50% patients feeling more educated about asthma triggers, more empowered to self-manage their asthma, and having fewer symptoms and home environmental triggers. We hypothesized that, these factors taken together, would minimize the impact of asthma on patients’ QOL.

## Methods

We evaluated the population served by the VHV program by asking a series of demographic- and housing-related questions during the first VHV. This included questions about the age of the participant, sex, race, homeownership status (homeowner or renter), level of income, and area of residence, among others. The full list of questions is available in **Supplement 1**. The LDH Institutional Review Board determined that the project met the criteria for exempt status (Nov 2022).

### Asthma symptom control

The first and last ACT scores were compared for patients in order to assess potential improvements in asthma control after participating in the BREATHE program. Initial and final scores (separated by approximately three months) were summarized for all patients who completed either test (unpaired comparison), as well as those who completed both (paired comparison), and testing using Mann-Whitney U-test. The 58 patients who completed both the initial and final ACT were also stratified by the number of VHV they had received (none, at least one, or all three) to test whether patients receiving the virtual visits experienced greater improvement in asthma control. Of note, in all data analyses, the ‘no VHV’ category included patients who did not qualify based on the pre-screening criteria as well as those who qualified but did not consent to receiving it. Non-parametric statistical tests were used to analyses data with non-normal distribution, as confirmed using the Shapiro-Wilk and Anderson-Darling tests.

### Environmental trigger education

Along with descriptive statistics, the percent patients who learned about at least one new asthma trigger after participating in the program was calculated. This was determined by tallying the number of patients who answered at least one question correctly in the post-test but not in the pre-test. Any increase in the post-test score (compared to the same person’s pre-test score) was also analyzed to identify the percent patients who showed overall greater knowledge of environmental asthma triggers. Patients showing improvements were then stratified by VHV status and compared using Fisher’s exact test to identify potential impact of VHV on knowledge gain.

### Patient attitudes and perceptions

Attitudes and perceptions regarding patients’ ability to self-manage asthma, following participation in the program, was assessed through a series of questions (listed below). Participants’ sense of empowerment to self-manage asthma after participating in the program was gauged based on their response to the question, “After participation in the program, do you feel more empowered to take control of your (or your child’s) asthma?” Answers ranged from ‘not at all’ (score=1) to ‘a lot’ (score=5), and a score of 3 or higher was considered a positive outcome. Perceptions regarding potential symptom reduction was measured based on responses to the question, “Since you participated in the program, how often have you (or your child) had asthma symptoms like coughing, wheezing or needing to use an inhaler?” Response options were ‘more often’, ‘less often’, ‘about the same’ and ‘none at all’.

Of these, ‘less often’ and ‘none at all’ were considered positive outcomes. To understand participants’ beliefs about environmental trigger reduction, we asked a true or false question: “Since participating in this program, I believe there are fewer asthma triggers in the home resulting from cleaning practices, pest control practices, smoking, etc.” Separately, we also asked patients about the specific environmental trigger reduction strategies they implemented. Patients’ perceptions about their level of education regarding asthma triggers was checked using the question: “How much did you learn about environmental asthma triggers by participating in this program?” Answers ranged from ‘not at all’ (score=1) to ‘a lot’ (score=5), and a score of 3 or higher was considered a positive outcome. Finally, participants were asked to score the impact that asthma has had on their QOL over the prior two weeks, first at enrollment, and again at the end of the program. Responses ranged from ‘none at all’ (score=1) to ‘a lot’ (score=5), and a score of 3 or lower was considered a positive outcome. The overall shift of percent clients experiencing the positive outcome (i.e., ‘none’, ‘not much’, or ‘a little’ impact) at the end of the intervention period, as compared to ‘quite a bit’ or ‘a lot’ in the beginning of the program, was tested using Fisher’s exact test. Each question was analyzed separately for this set of analyses, and the denominator consisted of the total number of patients who responded to the particular question. Stratification by VHV status was not possible for this analysis due to very few respondents in the “no VHV” group.

## Results

### Clinical, environmental and demographic characteristics of patients

Between March 2021-October 2022, 395 patients were referred, and 147 (37%) of them completed the enrollment requirements for the program. Of these, 112 (76%) qualified for the VHV, 52 of whom (46%) had consented and received at least one virtual visit as of October 2022. A majority of the 147 enrollees suffered from allergies (80%) and used a rescue inhaler ≥2 times in a typical week (69%) **(Table 1A)**. The most notable environmental characteristic present in homes was the use of strongly-scented cleaning agents and/or perfumed products (80%). Nearly half (47%) also had moderate or severe dust build-up in their homes **(Table 1B)**. About 61% of enrollees were from the three previously identified areas of concern: 25% and 23% from EBR and Caddo Parishes respectively, and 13% from the GNO Area. Based on detailed patient demographics, the visits primarily served non-Hispanic Black patients (76%), children (77%), renters (62%) and low-income families (90%) **(Table 1C)**. In terms of location, nearly half of VHV recipients (44%) were from EBR Parish.

### Asthma symptom control

Based on pre-vs. post-intervention ACT scores, asthma symptoms improved across all enrollees who completed either the initial and/or the final ACT, with a median increase of 2.39 points on the ACT (P=0.0102; **Table 2A**). Similar results were observed among the 58 participants who completed both the initial and final ACT with 62% patients showing improvement (P=0.0114; **Table 2B**). When stratified by virtual visit status, improvements in average and median ACT scores was observed for all groups; however, only patients who received all three VHV experienced statistically significant improvements (P=0.00634; n=25. Of note, 50% patients in both VHV groups started with an ACT score <19 (considered not well-controlled asthma) and ended with an ACT score >19, which is considered well-controlled asthma **(Table 2C)**.

**Table 2.** Comparison of pre- and post-intervention Asthma Control Test (ACT) scores among BREATHE patients.

| A) All ACT scores: |  |  |  |  |  |  |
| --- | --- | --- | --- | --- | --- | --- |
|  | Initial | Final | Change | P-value<sup>a</sup> |  |  |
| Average | 17.45 | 19.37 | +2.24 | 0.0102 |  |  |
| Standard deviation | 4.75 | 4.80 |  |  |  |  |
| Median | 17.50 | 20.00 | +2.39 |  |  |  |
| N | 86 | 62 |  |  |  |  |
| B) Paired ACT scores: |  |  |  |  |  |  |
|  | Initial | Final | Change | P-value<sup>a</sup> |  |  |
| Average | 17.33 | 19.57 | +2.24 | 0.0114 |  |  |
| Standard deviation | 5.05 | 4.71 |  |  |  |  |
| Median | 18.00 | 20.50 | +2.39 |  |  |  |
| N | 58 | 58 | 62% improved |  |  |  |
| C) Median ACT scores by Virtual Home Visit status: |  |  |  |  |  |  |
|  | Initial | Final | Change | % improved | N | P-value<sup>a</sup> |
| At least 1 VHV | 18.00 | 21.30 | +2.00 | 64% | 33 | 0.0524 |
| All 3 VHV | 18.00 | 21.30 | +3.00 | 76% | 25 | 0.0063 |
| No VHV | 15.00 | 19.00 | +4.00 | 60% | 25 | 0.0643 |

<sup>a</sup> P-value derived from Mann-Whitney U-test
N = number of patients who completed the survey
VHV = Virtual Home Visit

### Knowledge of environmental trigger management

There was no difference in the median knowledge test scores for the 50 enrollees who completed both instruments (83% [pre] vs. 88% [post]). However, slightly higher percent respondents scored in the 90-100% range on the post-intervention knowledge test (30% [pre] vs. 42% [post]). Of note, patients also showed greater awareness regarding the subtleties of environmental trigger reduction during the post-test, which suggested a deeper understanding of the subject material. For instance, on the true/false question about an open bathroom window removing moisture from the air, many patients considered the impact of indoor vs. outdoor humidity only during the post-test.

Comparing pre/post scores for each individual revealed that 86% of all respondents (n=43/50) learned about at least one new asthma trigger **(Figure 2A)**. There was no difference when stratified by VHV status; 88% respondents (n=22/25) who received VHV learned about at least one new environmental trigger compared to 84% respondents (n=21/25) who did not **(Figure 2B)**. About 52% respondents (n=26/50) had a higher post-intervention test score compared to their pre-test score **(Figure 2C)**. Here, there was a significant difference by VHV status (P=0.0465; Fisher’s exact test); 68% VHV recipients (n=17/25) showed an improved post-test score, compared to 36% of non-recipients (n=9/25) **(Figure 2D)**. This is consistent with the higher mode observed in the post-test scores of VHV recipients (88% [pre] vs. 96% [post]). Such increase was not observed for the ‘no VHV’ group whose mode remained at 92% both for the pre- and post-tests.

**Figure 2:**
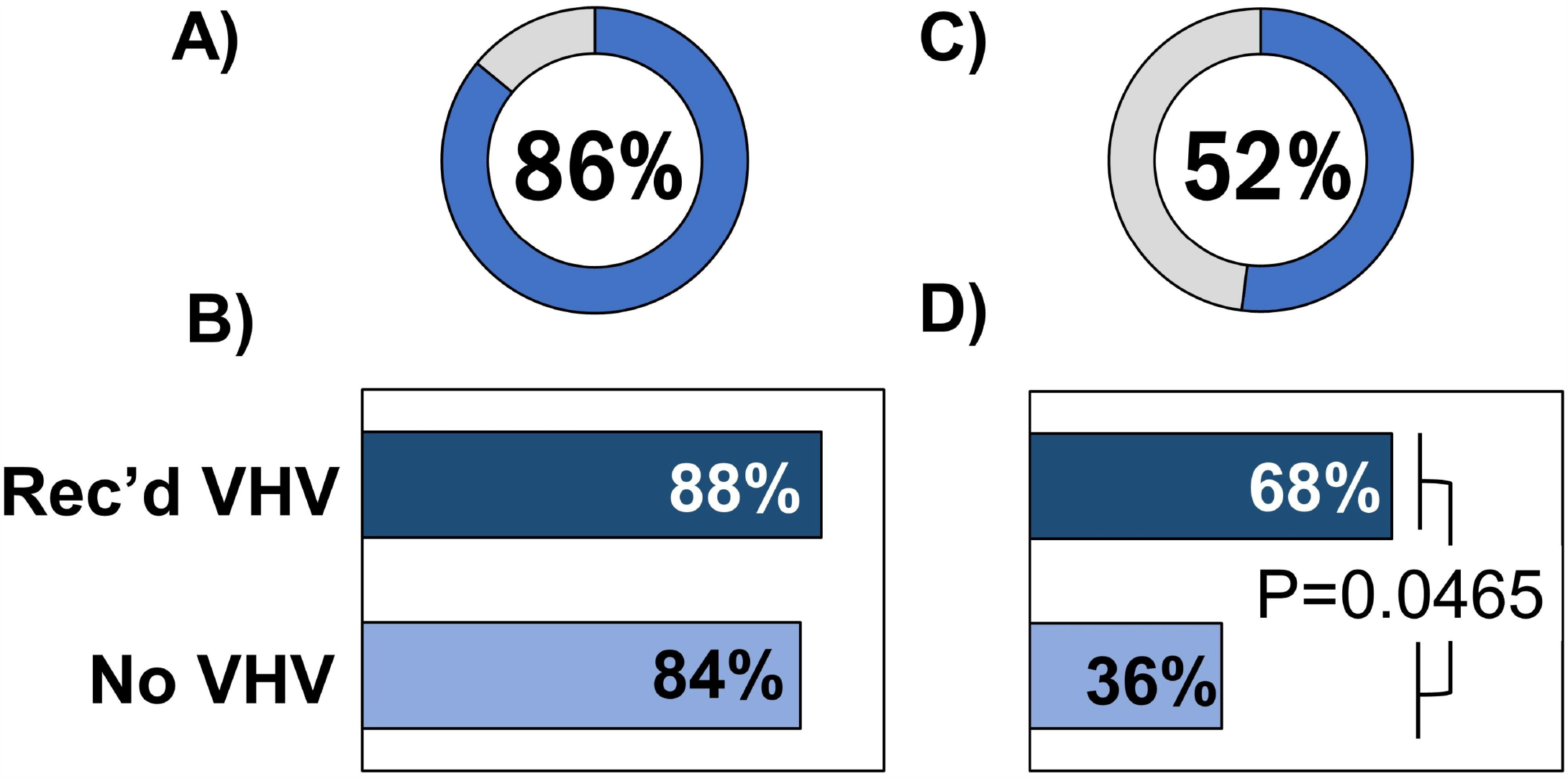
Knowledge gain among BREATHE participants. Percent respondents who learned about at least one new environmental trigger – overall **(A)** and stratified by VHV status **(B)**. Percent respondents with improved post-intervention knowledge test score – overall **(C)** and by VHV status **(D)**.

### Attitudes and perceptions about asthma self-management and impact of asthma on QOL

A majority of respondents reported having fewer symptoms (n=35/48; 73%) since participating in the program, and feeling more empowered (n=37/49; 75%) and educated (n=47/51; 94%) about self-managing their asthma **(Figure 3A)**. About 82% respondents also believed there to be fewer asthma triggers in the home (n=37/45), and reported implementing changes such as cleaning more frequently, using asthma-friendly cleaners, avoiding bleach, and ventilating the kitchen when cooking. At the end of the intervention period, there was also a statistically significant shift towards reduced impact from asthma on QOL **(Figure 3B)**. During enrollment, 33% of 106 total respondents reported the impact of asthma on their QOL being ‘a lot’. This number had reduced to 13% in the final survey, with 47 total respondents. Conversely, percent participants who reported ‘no’ impact of asthma on QOL rose from 16% during enrollment to 49% during the final survey.

**Figure 3:**
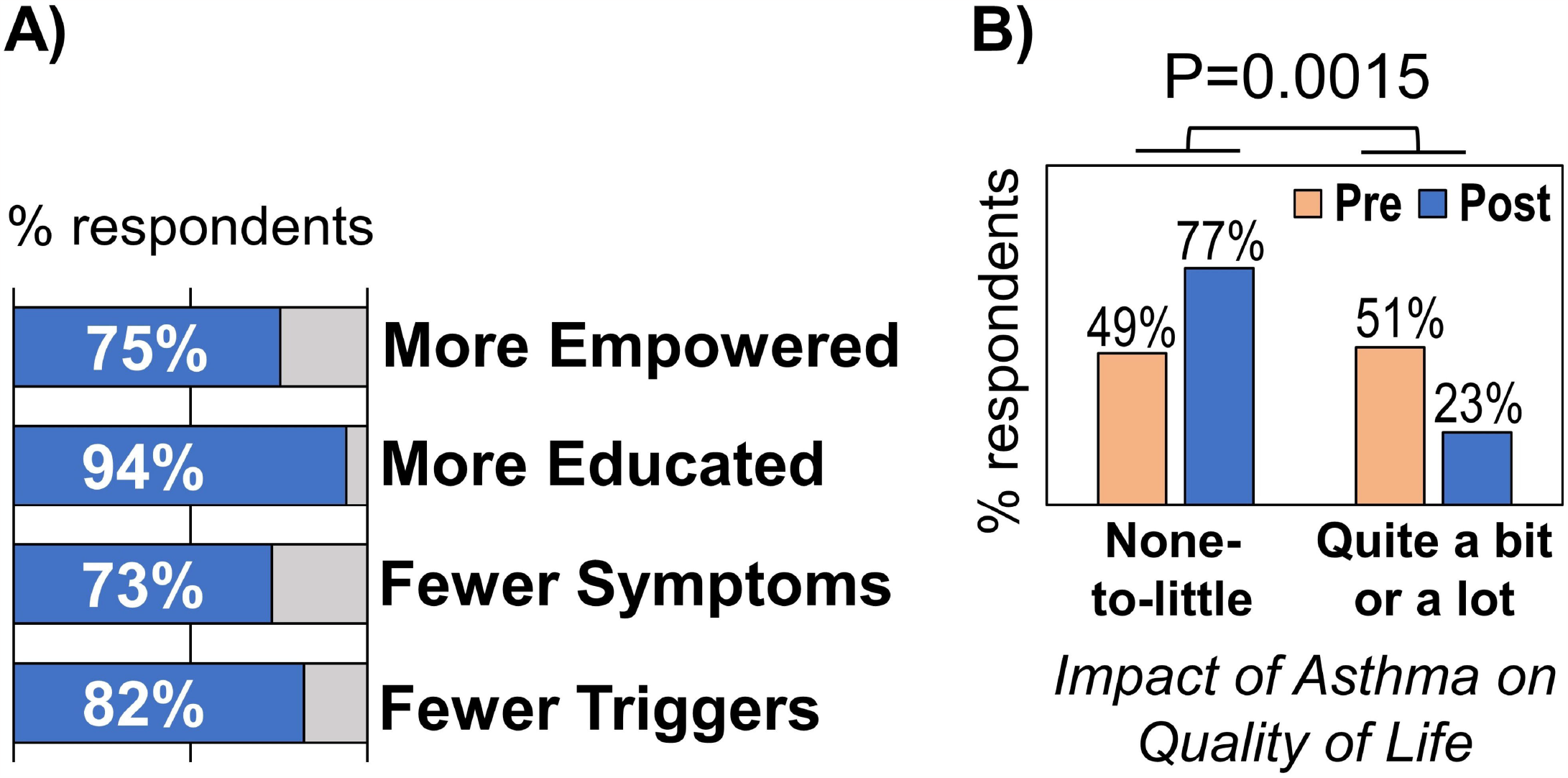
Perceptions regarding asthma self-management (A) and impact of asthma on quality of life (B).

## Discussion

At the start of the COVID-19 pandemic, many in-person public health programs made an attempt to implement virtual models that did not involve face-to-face contact. This was certainly the case for several asthma home visiting programs around the country. While much of this information was shared via websites, presentations and webinars,^12, 13, 14^ there is limited peer-reviewed literature on the practice and efficacy of these programs. To our knowledge, this is the first report to show distinct clinical improvements in asthma outcomes, using a validated measure (ACT scores), among patients who completed the VHV program **(Table 1)**. The data also show improvements in patients’ knowledge and perceptions regarding asthma self-management, and a reduced impact of asthma on quality of life among all BREATHE program participants **(Figure 2-3)**. As such, these data have important public health implications, not only for practitioners but also for policymakers and health plans as they consider coverage for virtual visits, especially for Medicaid patients.

### Lessons Learned

Lessons learned from the BREATHE VHV pilot are summarized below for public health practitioners:

1. Virtual visits can provide services to at-risk communities, and yield improvements in asthma symptoms. Thus, they can be a viable alternative to in-person home visits for asthma and Healthy Homes education. Indeed, the virtual model may even be the key to success with people who are uncomfortable with strangers visiting their homes because of privacy, safety or health concerns.
2. However, a better understanding is needed of the barriers faced by patients who qualified for VHV but did not consent to receiving them. If the barriers are related to technology, then partnering with internet-capable community centers may be helpful.
3. While all patients showed improvements, those receiving VHV showed greater improvements in knowledge of environmental trigger reduction. This suggests that additional time spent discussing the material with a trained asthma educator may be helpful for improving retention.
4. Further, if patients’ symptoms are severe, additional effort should be made to complete more than one VHV, as they may need more follow-up than patients whose asthma is more easily managed.
5. Expanding partnerships (especially with clinics and hospitals) to maximize enrollment, providing patients with additional, value-added wrap-around services, allowing patients to access the surveys in multiple ways (phone, email and/or text) and incorporating trust-building processes (eg: sharing photo of the asthma educator ahead of the VHV) can improve participation and retention. Services such as remediation for pests/mold (in coordination with landlords as needed) would strengthen the program’s impact as well.
6. The most common change reported by VHV recipients was change of cleaning agents. This was also the topic that patients showed most improvement on in the knowledge test. If the changes implemented as a result of knowledge gain played a role in improving their symptoms, such low-cost housekeeping modifications could be further promoted by providing patients with “green-cleaning” supplies.^15,16^
7. Given the disparities in asthma outcomes faced by low-income and Black children, the success of our program suggests that virtual models of asthma education can be a useful tool for reducing disparities and promoting health equity, especially in areas where (and times when) access to in-person home visits may be limited.

### Limitations

There are four limitations in this study. First, the virtual design did not allow for in-person medical and environmental evaluations, so the program relied on self-reported data. Second, because of the small sample size, we could not evaluate program impact stratified by demographic variables such as age or race. Third, the data collection mechanism was not designed to allow for certain comparisons, such as between patients who qualified for VHV but did not accept them and those who did, and any changes in healthcare utilization. Finally, there were an uneven number of respondents for the evaluation metrics as patients were not required to answer all the questions. Some of these limitations are being addressed in future iterations of the program, and the results will be described in future publications.

On the whole, we conclude that the BREATHE Virtual Home Visits program was successful in providing asthma education to disproportionately affected population. As such, it has an important public health impact.

## Data Availability

All data produced in the present study are available upon reasonable request to the authors via email at.

