## Supplemental File 1 for "Virtual Home Visits Reduce Asthma Burden in Low-Income, Black Communities amidst the COVID-19 Pandemic"

### Virtual Home Visit 1

Case ID ______________________

#### Initial Virtual Home Visit

Date and time of initial home visit ______________________

Name of Interviewer
- ◯ Tracy Marquette

Other Staff assigned ______________________

Status of Home Visit
- ◯ Successful
- ◯ Unsuccessful

If unsuccessful, reason:
- ◯ client requested to reschedule
- ◯ client refused services (cancelled)
- ◯ Client unavailable at home (no show)
- ◯ other

Status
- ◯ Visit Complete
- ◯ No Visit

Reason for no visit
- ◯ reschedule
- ◯ moved
- ◯ dismiss landlord issues
- ◯ dismiss insurance issues
- ◯ other

other reason for no visit ______________________

#### Client Contact Information

Do I have your consent to carry out this home visit, to ask you questions about your home and your child's asthma for the purpose of providing educational materials that may reduce the environmental triggers in your home? Yes or NO
- ◯ Yes
- ◯ No

First Name ______________________

Last Name ______________________

Primary number ______________________

Alternative Number

______________________________

Address

______________________________

Patient Name

______________________________
(this is who suffers from asthma )

Patient Age

______________________________

#### Household Information

Female Head of Household

◯ No
◯ Yes

Total number of persons living in household

______________________________

What is the approximate household annual income?

______________________________

Do you like the neighborhood?

◯ Yes
◯ No

Do you think this is someplace you would like to stay for a while?

◯ Yes
◯ No

Please explain why or why not you would want to stay in in this neighborhood.

______________________________

How many times in the last 12 months have you changed residences?

◯ none
◯ 1 time
◯ 2 times
◯ 3 times
◯ 4 times
◯ 5 or more times

Do you rent or own?

◯ Rent
◯ Own

Do you have a mortgage on the home?

◯ Yes
◯ No

How much is your rent or mortgage per month?

______________________________

Are you currently late on your rent or mortgage payments?

◯ Yes
◯ No

| | |
|---|---|
| If you are currently late on rent or mortgage payment, what is the cause? | ☐ Loss of Income<br>☐ Reduction in Income<br>☐ Medical Issues<br>☐ Increase in Expenses<br>☐ Divorce/Separation<br>☐ Death of Family Member<br>☐ Increase in Mortgage Payment<br>☐ Budget Management Issues<br>☐ Business Venture Failure<br>☐ Other |
| Do you have homeowner's or renter's insurance | ◯ Yes<br>◯ No |
| Do your receive Section 8 or other housing assistance? | ◯ Yes<br>◯ No |
| if yes, what program? | ______________________ |
| Do you live in public housing? | ◯ Yes<br>◯ No |

#### Asthma Questions

| | |
|---|---|
| Name of patient with asthma | ______________________ |
| Gender of patient | ◯ male<br>◯ female |
| date of birth | ______________________ |
| age of patient at time of visit | ______________________ |
| Does the person with asthma also have a disability? | ◯ No<br>◯ Yes |
| Is this person of Hispanic, Latino of Spanish origin? | ◯ No<br>◯ Yes |
| What is this person's race? | ☐ Black<br>☐ White<br>☐ American Indian or Alaskan Native<br>☐ Asian<br>☐ Pacific Islander<br>☐ Other |
| If Other specify race/ethnicity | ______________________ |

| | |
|---|---|
| What does 'successful asthma management' mean for you?<br><br>For example, 'successful asthma management' could mean you can go dancing again, or do more gardening (or other hobby), or simply feel less worn out during the course of your daily activities. | ___________________________ |
| How are your/your child's asthma symptoms right now? | ◯ out of control<br>◯ poorly controlled<br>◯ somewhat controlled<br>◯ well controlled<br>◯ I don't know |
| What triggers your/your child's asthma? select all that apply | ☐ animals/pets<br>☐ cleaning supplies (bleach, detergents, etc)<br>☐ dust mites<br>☐ exercise/physical activity<br>☐ food allergies<br>☐ illnesses (cold, respiratory infections, etc)<br>☐ mold<br>☐ strong smells (perfumes, fragrances, etc)<br>☐ pollen<br>☐ pollution<br>☐ pests/rodents (mice, rats, cockroaches, etc)<br>☐ tobacco smoke<br>☐ weather<br>☐ intense emotions/stress<br>☐ other<br>☐ don't know/uncertain<br>(Use the Indoor Air Pollution and EPA checklist front page for triggers.) |
| If other please explain | ___________________________ |
| Did you know there are "asthma-friendly" ways to clean surfaces in a way that kills the COVID virus? | ◯ Yes<br>◯ No |
| Below are some "asthma-friendly" ways to kill the COVID virus. Which, if any of these, can work for your household? | ☐ Use low-odor disinfectants like Ethyl Alcohol (rubbing alcohol) or up to 3% Hydrogen Peroxide and not products meant for industrial or hospital use.<br>☐ Have the asthmatic person stay in another room when cleaners or disinfectants are being used and right after their use.<br>☐ Limit use of chemicals that can trigger asthma attacks, such as bleach or ammonium compounds, and do not mix the or use them in enclosed spaces.<br>☐ Use only cleaning products you must use. Some surfaces and objects that are seldom touched may need to be cleaned only with soap and water.<br>(Reference to the Green Cleaning handout) |
| Have you noticed anything that seems to trigger your/your child's asthma? | ___________________________ |
| Have/has you/your child ever had an allergy test? | ◯ Yes<br>◯ No<br>(mention mold and allergic rhinitis handout) |

| | |
|---|---|
| If Yes, what were the results? | ______________________ |
| Is there a secondary household where you/your child spends time regularly? (Grandparents' house, other parent's house, etc) | ◯ Yes<br>◯ No |
| If yes, please explain any changes in asthma symptoms with the change in environment. | ______________________ |
| How long have you lived at this address? | ◯ < 3 months<br>◯ 3-6 months<br>◯ 6 months-1 year<br>◯ 1-3 years<br>◯ >3 years |
| Has your/your child's asthma symptoms changes since moving to this address? | ◯ Yes<br>◯ No |
| If yes, how? | ______________________ |
| When did you/your child last have symptoms? | ______________________ |

#### Asthma Health Care Utilization: Past 3 Months

| | |
|---|---|
| How would you rate your/your child's asthma during the past 3 months? | ◯ out of control<br>◯ poorly controlled<br>◯ somewhat controlled<br>◯ well controlled<br>◯ I don't know |
| In the past 3 months how many days of work/school/daycare have/has you/your child missed due to asthma? | ◯ 0<br>◯ 1<br>◯ 2<br>◯ 3<br>◯ 4<br>◯ 5<br>◯ greater than 5 |
| In the past 3 months, how often did your/your child's asthma keep you from getting as much work done? (at home, work, or school) | ◯ 0<br>◯ 1<br>◯ 2<br>◯ 3<br>◯ 4<br>◯ 5<br>◯ greater than 5 |
| In the last 3 months, how many days have/has you/the child been working harder to breathe? | ◯ 0<br>◯ 1<br>◯ 2<br>◯ 3<br>◯ 4<br>◯ 5<br>◯ greater than 5 |

| | |
|---|---|
| In the last 3 months, how many days did asthma symptoms wake you/your child up in the middle of the night or earlier than usual in the morning? | ◯ 0<br>◯ 1<br>◯ 2<br>◯ 3<br>◯ 4<br>◯ 5<br>◯ greater than 5 |
| In the past 3 months, how many times has your/your child's asthma caused you to call your doctor? | ◯ 0<br>◯ 1<br>◯ 2<br>◯ 3<br>◯ 4<br>◯ 5<br>◯ greater than 5 |
| In the past 3 months, how many times has your/your child's asthma caused you to go to the Emergency Room or Urgent Care Clinic? | ◯ 0<br>◯ 1<br>◯ 2<br>◯ 3<br>◯ 4<br>◯ 5<br>◯ greater than 5 |
| In the past 3 months, besides emergency room/urgent care visits, how many times has your/your child's asthma caused you to go to your doctor's office or clinic for worsening of symptoms? | ◯ 0<br>◯ 1<br>◯ 2<br>◯ 3<br>◯ 4<br>◯ 5<br>◯ greater than 5 |
| In the past 3 months, how many times have/has you/your child been admitted overnight in a hospital due to asthma? | ◯ 0<br>◯ 1<br>◯ 2<br>◯ 3<br>◯ 4<br>◯ 5<br>◯ greater than 5 |
| How many times have/has you/your child EVER in his/her lifetime been to the ER/UCC as a result of asthma? | ◯ 0<br>◯ 1<br>◯ 2<br>◯ 3<br>◯ 4<br>◯ 5<br>◯ < 5 |
| In the past month how many days did your child's asthma keep you from performing normal daily activities (at home, work or school)? | ______________________ |

#### How do the following affect your/your child's asthma symptoms?

Humidity:

◯ Improves symptoms
◯ No change
◯ Makes symptoms worse

Air Conditioning

◯ Improves symptoms
◯ No change
◯ Makes symptoms worse

#### Medical Information

#### Here is where refering to the OLOL Asthma Handbook may be the most helpful

Who is the primary care physician?

______________________________

phone number

______________________________

PCP Address

______________________________

How long have/has you/your child been seeing this doctor?

______________________________

Additional asthma or allergy related specialists:

______________________________

Do you use a Controller Medication?

◯ Yes
◯ No

What type of medication is it?

◯ Inhaler
◯ Liquid
◯ Nasal Spray
◯ Pill
◯ Other

What is the Brand of the medication?

______________________________

How are you meant to take the medication?

______________________________

Date Filled:

______________________________

Expiration Date:

______________________________

Number of Refills:

______________________________

| | |
|---|---|
| Are you taking this medication as directed (right dose and on time)? | ◯ Yes--always!<br>◯ Yes--most of the time<br>◯ Yes, but sometimes I don't/can't<br>◯ Most of the time I don't/ can't<br>◯ I almost never do/ can |
| Completed education on Controller medication? | ◯ Yes<br>◯ No |
| Do you have a quick relief medication? | ◯ Yes<br>◯ No |
| What type of medication is it? | ◯ Inhaler<br>◯ Liquid<br>◯ Nasal Pray<br>◯ Pill<br>◯ Other |
| What is the brand of the medication? | ______________________________ |
| How are you meant to take the medication? | ______________________________ |
| Date Filled: | ______________________________ |
| Expiration Date: | ______________________________ |
| Number of Refills: | ______________________________ |
| Are you taking this medication as directed (right dose and on time)? | ◯ Yes--always!<br>◯ Yes--most of the time<br>◯ Yes, but sometimes I don't/can't<br>◯ Most of the time I don't/can't<br>◯ I almost neve do/can |
| Completed education on Quick Relief Meds? | ◯ Yes<br>◯ No |
| Are you taking any other medications? | ◯ Yes<br>◯ No |
| Please list all other medications | ______________________________ |
| In the past 6 months how many times have/has you/your child been prescribed Prednisone? | ______________________________ |
| Have you/ your child ever been intubated due to complications with asthma? | ◯ Yes<br>◯ No |
| if yes, when was this? | ______________________________ |

| | |
|---|---|
| How would you rate the effectiveness of your/your child's medications? (5=Very Effective and 0=Not Effective) | ◯ 0<br>◯ 1<br>◯ 2<br>◯ 3<br>◯ 4<br>◯ 5 |
| Besides medication what else have you found is helpful to your/your child's asthma? | ________________________ |
| Who administers the child's meds and treatments most often? | ________________________ |
| Does your child have a prescription (rescue inhaler) at school? | ◯ Yes<br>◯ No |
| Does your child have a spacer at school? | ◯ Yes<br>◯ No |
| How frequently does the patient use the spacer with his/her controller medication? | ◯ Never<br>◯ Rarely<br>◯ Sometimes<br>◯ Very Often<br>◯ Always |
| How frequently does the paient use the spacer with his/her rescue inhaler? | ◯ Never<br>◯ Rarely<br>◯ Sometimes<br>◯ Very Often<br>◯ Always |
| Notes on Medication Utilization | ________________________ |
| Does you/your child use a peak flow meter? | ◯ Yes<br>◯ No |
| Has your health care provider/doctor/nurse every given you an asthma action plan? | ◯ Yes<br>◯ No |
| If yes, when? | ________________________ |
| Does the asthma patient have health insurance? | ◯ Yes<br>◯ No |
| What kind of insurance? | ◯ Medicaid<br>◯ Private Insurance<br>◯ Amerigroup<br>◯ United Health Care<br>◯ Blue Cross Blue Shield<br>◯ Aetna<br>◯ Other |

**Level of Intervention Questions**

**The EPA Checklist should be explained here. You do not need to collect the answers from the whole checklist just what is listed below.**

| | |
|---|---|
| Do you have a working vacuum? | ◯ Yes<br>◯ No |
| What kind (brand)? | ______________________ |
| Does it have a HEPA filter? | ◯ Yes<br>◯ No |
| Does the home have wall to wall carpet? | ◯ Yes<br>◯ No |
| Do you have a kitchen hood above your stove that vents to the outside? | ◯ Yes<br>◯ No |
| Do you have a kitchen exhaust fan or a window in the kitchen that opens? | ◯ Yes<br>◯ No |
| Do you use a dust mite allergy mattress cover on your bed? | ◯ Yes<br>◯ No |
| What size mattress do you have? | ◯ single<br>◯ twin<br>◯ full<br>◯ queen<br>◯ king<br>◯ other |
| If other, please specify: | ______________________ |
| Are you aware of Air Quality Index (AQI) and the Air Quality "Alert" Days from EPA? | ◯ Yes<br>◯ No |
| How can you protect yourself when the outdoor air quality is poor? | ◯ Avoid being outside in the afternoon & early evening. That's when air pollution levels are usually highest<br>◯ Do less physical activity. Physical activity increases the amount of air you breathe.<br>◯ Keep doors and windows closed to keep harmful air out of your home<br>◯ Change air filters and run air conditioning in "recirculating mode"<br>◯ All of the above<br>(This is where you should have them look at the Outdoor AQ & Asthma handout) |
| AQI:<br>Demonstration of how to use the app with the AQI value of the day.<br>Make sure the family has signed up for the text alerts and has the app downloaded | ◯ Yes<br>◯ No |

| | |
|---|---|
| SEET hotline<br>Please make sure that the family has the SEET hotline number<br>(888) 293-7020<br>They may use this number to contact us for any concerns they have about their indoor health. | ◯ Yes<br>◯ No<br>(Mention asthma class at OLOL) |
| What school does your child attend?<br>Schools name and address | ____________________ |
| Date of Virtual Home visit 2<br>Schedule the next visit now with the patient | ____________________ |
