## Supplemental File 2 for "Virtual Home Visits Reduce Asthma Burden in Low-Income, Black Communities amidst the COVID-19 Pandemic"

### Program Evaluation

Thank you for participating in this BREATHE Asthma Outreach Program. We hope it was helpful to you and your family. We would like to ask you a few questions – how you felt about the program, what you thought about our work, and what you might do with the information and materials you received. Your answers will help us improve our outreach and reach other families.

Date: ____________________

Name of participate answering questions ____________________

#### Questions for all participants:

How has your (or your child's) asthma symptoms been during the past month? ____________________

After participation in this program, do you feel more empowered to take control of your (or your child's) asthma?
- ○ Not at all ○ Not much
- ○ Maybe a little ○ Quite a bit
- ○ Yes, Definitely!

How much did you learn about environmental asthma triggers by participating in this program?
- ○ Not at all
- ○ Not much
- ○ Maybe a little
- ○ Quite a bit
- ○ I learned a lot !

After participating in this program, I know where I can get personalized help if I have a question about indoor environmental quality and Healthy Homes.
- ○ Yes
- ○ No

During your first home visit, you discussed what successful asthma management meant personally to you. Do you feel this program has helped make that goal more achievable for you?
- ○ Yes, definitely!
- ○ Yes, somewhat
- ○ Maybe a little
- ○ Maybe, but not much
- ○ No, not at all

Did you sign up for Outdoor Air Quality email alerts through EPA's EnviroFlash or Louisiana's Dept. of Environmental Quality (LDEQ)?
- ○ Yes
- ○ No--I didn't even know that was an option!
- ○ No--I knew about it but did not sign up

If you knew about it but did not sign up, why not? ____________________

How helpful were the education materials about asthma triggers and cleaning methods that you received?
- ○ Not at all helpful
- ○ Not helpful
- ○ I'm not sure
- ○ Helpful
- ○ Very helpful

Do you think you will continue to use the information and household practices you learned about?
- ○ Yes
- ○ No

| | |
|---|---|
| Since you found out about it, how often have you used the resources from EPA or LDEQ to check the outdoor air quality in your area? | ◯ I use it all the time!<br>◯ Every now and then<br>◯ maybe once or twice<br>◯ Never --I didn't even know that was an option!<br>◯ Never--I knew about it but never used it |
| If you knew about it, but never checked the outdoor air quality in your area, why not? | __________________________________ |
| Since you participated in this program, how often have you (or your child) had asthma symptoms like coughing and wheezing, or needing to use an inhaler? | ◯ More often<br>◯ Less often<br>◯ About the same amount<br>◯ No asthma symptoms since! |
| After participating in this program, did you make any changes in how you clean and maintain your home? | ◯ Yes, a big change<br>◯ A few things, a small change<br>◯ No, not really |
| If you made any changes, please describe them briefly. | __________________________________ |
| If you have NOT made any changes yet (or even just a few changes), do you plan to make any (more) changes later based on what you learned? | ◯ Yes<br>◯ No |
| TRUE or FALSE: Since participating in this program, I believe there are fewer asthma triggers in the home resulting from cleaning practices, pest control practices, smoking, etc. | ◯ True<br>◯ False |
| How much do you feel ASTHMA has affected your quality of life in the past two weeks? Consider, for instance, the number of missed work/school days due to asthma, lost productivity, how often you felt limited in your activities (e.g., exercising, running, gardening, cleaning, etc.), and/or how often your mental health has been affected due to asthma (e.g., if you felt sad, depressed, "left out", "different", anxious or frustrated because of asthma). | ◯ Not at all<br>◯ Not much<br>◯ Maybe a little<br>◯ Quite a bit<br>◯ A lot |

#### What did you think of the virtual home visits you had where asthma management and Healthy Homes were discussed?

| | Yes, I agree | No, I disagree | N/A |
|---|---|---|---|
| I liked working with the Our Lady of the Lake Children's Hospital and Louisiana Health Department | ◯ | ◯ | ◯ |
| I felt that the visits took too long | ◯ | ◯ | ◯ |
| I liked the information I was given at the home visits | ◯ | ◯ | ◯ |

| | | | |
|---|---|---|---|
| I felt that the home visits/questionnaire invaded our privacy | ◯ | ◯ | ◯ |
| I liked the education provided during the virtual visits | ◯ | ◯ | ◯ |
| I felt that the information didn't apply to me and my family | ◯ | ◯ | ◯ |
| I would like to learn more about managing asthma | ◯ | ◯ | ◯ |

---

How many days of work days and/or school days (choose one or both) have you missed in the past 4 weeks because of your or your child's asthma?

- ☐ 1 day or less of school
- ☐ 2-5 days of school
- ☐ more than 5 days of school
- ☐ 1 day or less of work
- ☐ 2-5 days of work
- ☐ more than 5 days of work

---

Choose all that apply:  Is/are there a particular place(s) that you find your asthma symptoms have been worse in the past 4 weeks?

- ☐ Home
- ☐ Workplace
- ☐ School
- ☐ Other

---

If other, please describe

__________________________________

---

What else would have made this program more useful to you?

__________________________________

---

Is there anything else you would like to tell us about your asthma virtual home visits?

__________________________________________
