## Supplemental FIle 3 for "Virtual Home Visits Reduce Asthma Burden in Low-Income, Black Communities amidst the COVID-19 Pandemic"

### Environmental Asthma Trigger Knowledge Test

#### Pre-Knowledge Check / Post-Knowledge Check

Today's Date

______________________________

#### For each of the following indicate whether you think it can cause ("trigger") an asthma attack for ANYONE:

| | Yes | No | I'm Not Sure |
|---|---|---|---|
| Tobacco smoke | ○ | ○ | ○ |
| Floors made from concrete, tile, or linoleum | ○ | ○ | ○ |
| Animal dander from furry animals (like dogs and cats) | ○ | ○ | ○ |
| Cockroaches | ○ | ○ | ○ |
| Snakes or reptiles | ○ | ○ | ○ |
| Mold and mildew | ○ | ○ | ○ |
| Smoke from a fire, fire pit, or fireplace | ○ | ○ | ○ |
| Cleaning chemicals with strong odors | ○ | ○ | ○ |
| Dust mites | ○ | ○ | ○ |
| Pollen from plants and trees | ○ | ○ | ○ |
| Outdoor pollutants like ozone, particulate matter | ○ | ○ | ○ |

#### Are the following statements true of not?

| | Yes, True | No, Not True | I'm Not Sure |
|---|---|---|---|
| Vaccuming or wet mopping at home at least once a week can help prevent asthma attacks for people | ○ | ○ | ○ |
| Getting rid of food sources and breeding places is important to control and prevent pests | ○ | ○ | ○ |
| Using a kitchen exhaust fan or keeping kitchen & bathroom windows slightly open will remove moisture from the air | ○ | ○ | ○ |

| | | | |
|---|---|---|---|
| A water leak in the home can result in mold or mildew | ○ | ○ | ○ |
| Leaky plumbing and standing water attracts pests | ○ | ○ | ○ |
| Mold and mildew indoors can make asthma worse | ○ | ○ | ○ |
| Covering a pillow and mattress with an airtight cover can make asthma worse | ○ | ○ | ○ |
| Pests like cockroaches don't make any difference to a person with asthma | ○ | ○ | ○ |
| The stronger a cleaning product is (like full strength bleach), the better it is for cleaning your house and prevent asthma attacks | ○ | ○ | ○ |
| In addition to taking the right medication, there are other things people can do to prevent asthma attacks | ○ | ○ | ○ |
| Hydrogen peroxide (up to 3%) and Ethanol (ethyl alcohol) are "asthma-friendly" disinfectants that kill the virus causing COVID-19 | ○ | ○ | ○ |
| You can find out how good (or not!) your outdoor air quality is by entering your ZIP code on the EPA's AirNOW website. | ○ | ○ | ○ |
| Reducing physical activity, and staying indoors with the doors and windows closed, may help prevent an asthma attack when the outdoor air quality is poor. | ○ | ○ | ○ |
