## Supplemental File 4 for "Virtual Home Visits Reduce Asthma Burden in Low-Income, Black Communities amidst the COVID-19 Pandemic"

### Asthma Control Test

| | |
|---|---|
| Date | ______________________ |
| How old is subject? | ◯ 4-11<br>◯ 12 or older |

#### ACT (ages 4-11)

| | |
|---|---|
| 1. How was your Asthma Today? | ◯ 0-Very Bad<br>◯ 1-Bad<br>◯ 2-Good<br>◯ 3-Very Good |
| 2. How much of a problem is your asthma when you run, exercise or play sports? | ◯ 0-It's a big problem, I can't do what I want to do.<br>◯ 1-It's a problem and I don't like it.<br>◯ 2-It's a problem but it's okay.<br>◯ 3-It's not a problem. |
| 3. Do you cough because of your asthma? | ◯ 0-Yes, all the time.<br>◯ 1-Yes, most of the time.<br>◯ 2-Yes, some of the time.<br>◯ 3-No, none of the time. |
| 4. Do you wake up during the night because of your asthma? | ◯ 0-Yes, all of the time.<br>◯ 1-Yes, most of the time.<br>◯ 2-Yes, some of the time.<br>◯ 3-No, none of the time. |
| 5. During the last 4 weeks, on average, how many days per month did your child have any daytime asthma symptoms? | ◯ 5 - Not at all<br>◯ 4 - 1-3 days/mo<br>◯ 3 - 4-10days/mo<br>◯ 2 - 11-18 days/mo<br>◯ 1 - 19-24 days/mo<br>◯ 0 - Everyday |
| 6. During the last 4 weeks, on average, how many days per month did your child wheeze during the day because of asthma? | ◯ 5 - Not at all<br>◯ 4 - 1-3 days/mo<br>◯ 3 - 4-10days/mo<br>◯ 2 - 11-18 days/mo<br>◯ 1 - 19-24 days/mo<br>◯ 0 - Everyday |
| 7. During the last 4 weeks, on average, how many days per month did your child wake up during the night because of asthma? | ◯ 5 - Not at all<br>◯ 4 - 1-3 days/mo<br>◯ 3 - 4-10days/mo<br>◯ 2 - 11-18 days/mo<br>◯ 1 - 19-24 days/mo<br>◯ 0 - Everyday |

| **ACT (ages 12 and older)** | |
|---|---|
| 1. In the past 4 weeks, how much of the time did your asthma keep you from getting as much done at work, school or at home? | ◯ 1 - All of the time<br>◯ 2 - Most of the time<br>◯ 3 - Some of the time<br>◯ 4 - A little of the time<br>◯ 5 - None of the time |
| 2. During the past 4 weeks, how often have you had shortness of breath? | ◯ 5 - Not at all<br>◯ 4 - Once or twice a week<br>◯ 3 - 3 to 6 times a week<br>◯ 2 - Once a day<br>◯ 1 - More than once a day |
| 3. During the past 4 weeks, how often did your asthma symptoms (wheezing, coughing, shortness of breath, chest tightness, or pain) wake you up at night or earlier than usual in the morning? | ◯ 5 - Not at all<br>◯ 4 - Once or twice<br>◯ 3 - Once a week<br>◯ 2 - 2 or 3 nights a week<br>◯ 1 - 4 or more nights a week |
| 4. During the past 4 weeks, how often have you used your rescue inhaler or nebulizer medication (such as albuterol)? | ◯ 5 - Not at all<br>◯ 4 - Once a week or less<br>◯ 3 - 2 or 3 times per week<br>◯ 2 - 1 or 2 times per day<br>◯ 1 - 3 or more times per day |
| 5. How would you rate your asthma control during the past 4 weeks? | ◯ 5 - Completely Controlled<br>◯ 4 - Well Controlled<br>◯ 3 - Somewhat Controlled<br>◯ 2 - Poorly Controlled<br>◯ 1 - Not Controlled at all |
| ACT Score:<br>***(Ages 4-11)*** | ______________________ |
| ACT Score:<br>***(Ages 12 and older)*** | ______________________ |
